# The childhood cancer registry in Switzerland: methods and results in 2025

**DOI:** 10.1101/2025.10.26.25338836

**Authors:** Grit Sommer, Marina Haller, Sophia della Valle, Christina Çinar-Kaufmann, Ursula M. Kuehnel, Christina Schindera, Fabiën N. Belle, Nicolas Waespe, Ben D. Spycher, Claudia E. Kuehni

## Abstract

**Aims:** Nationwide registration of childhood cancers in Switzerland began in 1976. With the implementation of the Federal Law on Cancer Registration in 2020, cancer registration became compulsory, and methods were standardized nationwide across childhood and adult cancers. This paper describes the objectives and methods of the Childhood Cancer Registry (ChCR) as of 2025 and presents recent data on incidence, mortality and survival of childhood and adolescent cancers in Switzerland.

**Methods:** The ChCR records all primary cancers diagnosed in children and adolescents aged 0–19 years who live in Switzerland. Case reporting is mandatory for physicians, hospitals, and pathology laboratories. Eligible diagnoses include all malignant tumors, tumors of uncertain/unknown behavior, benign central nervous system tumors, histiocytosis, and selected other conditions. Cases are coded primarily according to ICD-10, ICD-O, the International Classification of Childhood Cancer (ICCC), and the Toronto Childhood Cancer Staging Guidelines. The registry collaborates closely with the Swiss Paediatric Oncology Group (SPOG), Cantonal Cancer Registries (CCRs), the National Agency for Cancer Registration (NACR), and the Federal Statistical Office (FSO). We calculated incidence and mortality rates for patients diagnosed between 2014 and 2023 using FSO population statistics and estimated 1-, 5- and 10-year observed survival using the Kaplan-Meier method with the period approach.

**Results:** From 1976 to 2023, 12,665 primary cancers diagnosed in individuals aged 0–19 years were registered. Between 2014 and 2023, 2,441 new cases were diagnosed among children (0–14 years) and an estimated 1,214 among adolescents (15–19 years), adjusting for incomplete regional coverage in adolescents before 2020. In children, the most frequent diagnostic groups were CNS tumors (653, 27%), leukemias (641, 26%) and lymphomas (275, 11%), followed by neuroblastoma, soft tissue sarcomas and other carcinomas and melanomas (all 6%), kidney tumors (5%) and bone tumors (4%). In adolescents, the most common diagnoses were lymphomas (287, 24%), carcinomas and melanomas (277, 23%), and CNS tumors (213, 18%), followed by leukemias (12%) and germ cell tumors (10%). Age-standardized annual incidence per million (standardized to the European Standard Population 2013) was 189 (95% CI 182– 197) in children aged 0–14, and 282 (95% CI 265–299) in those aged 15–19. Overall, five-year survival was 89% for children and 90% for adolescents, with variation across ICCC-3 diagnostic groups.

**Conclusions:** With mandatory reporting and strong collaboration among national stakeholders, the ChCR has evolved into a comprehensive and reliable data source. Its high completeness and long-term follow-up make it a critical resource for epidemiological and clinical research on childhood and adolescent cancers in Switzerland.

## Introduction

Although childhood cancers are rare, they remain the leading cause of disease-related death among children and adolescents aged 1 to 19 years [1]. Survivors commonly face long-term health complications caused by the cancer itself or its intensive treatment [2]. As with other rare diseases, centralized data collection is essential for generating robust evidence that can help reduce incidence and improve treatment outcomes, survival, and quality of life. Dedicated, nationwide childhood cancer registries have been founded in many countries including Germany, Switzerland, the United Kingdom, France (with separate registries for hematological and solid tumors), Hungary, and others [3–7]. These registries enable national monitoring of cancer incidence and survival, and contribute to international studies where large-scale data enable meaningful analyses of rare cancer outcomes.

The childhood cancer registration in Switzerland developed in three main phases. The first began in 1976 — at the time only the cantons Geneva and Vaud/Neuchâtel had (general) cancer registries in place — when the Swiss Paediatric Oncology Group (SPOG) launched a national registry for children treated in clinical studies. By 1981, the registry expanded to include patients not enrolled in studies. It documented all cases treated in the nine SPOG clinics, regardless of whether the diagnosis had occurred in Switzerland. This covered nearly all cancers occurring in children up to age 15, and cancers among older adolescents that were treated in pediatric settings. In 1992, the registry started to record long-term outcomes, including relapses, death and late adverse effects of treatment.

The second phase began in 2003, when the registry moved to the Institute of Social and Preventive Medicine (ISPM) at the University of Bern and became the Swiss Childhood Cancer Registry (SCCR). There, it transitioned into a population-based registry covering all cancer patients diagnosed or treated in Switzerland up to age 20. The registry renewed its database, validated and coded diagnoses according to current classification systems, and cross-checked case completeness with hospital records and cantonal registries, both prospectively and retrospectively back to 1976. Research projects that used the data contributed financial support to registry activities, alongside funding from the SPOG, patient organizations, the Swiss Cancer League, and later, federal and cantonal contributions. The former SCCR has been described in detail [8].

The third phase began with the implementation of the Cancer Registration Act (CRA) in Switzerland in 2020, when cancer registration became compulsory nationwide [9]. Registration of childhood cancers (ages 0–19) remained centrally organized at the national level, because of the low case numbers and the need for specialized expertise, while adult cancer registration (ages 20+) continued at the cantonal level. The SCCR has been renamed to Childhood Cancer Registry (ChCR) and is now a federally funded entity, with operations mandated to the Institute of Social and Preventive Medicine (ISPM) at the University of Bern. Its inclusion criteria were adapted to register only incident cancers in children and adolescents of the permanent resident population of Switzerland at the time of diagnosis, excluding foreign residents. Data on all eligible patients diagnosed before 2020 were incorporated from the former SCCR data base.

Under the new legal framework, the ChCRs purpose is restricted to data collection and monitoring. It makes data available for research under strict conditions but does not conduct research independently. The registry has harmonized data collection and validation processes in close collaboration with Cantonal Cancer Registries (CCRs), the National Agency for Cancer Registration (NACR), and the Federal Statistical Office (FSO). The ChCR is responsible for validation, data management, quality control, handling third-party data requests, routine statistical reporting, and international collaboration for cancer in individuals under 20 years, thus performing the analogous tasks as the NACR for cancers diagnosed at older ages.

This publication describes the functioning of the ChCR since the implementation of the Cancer Registration Act in 2020, outlines changes from earlier registry phases, presents current data on registered cases, incidence, mortality, and survival, and provides an overview of research activities supported by the registry.

## Methods

### Role of the Childhood Cancer Registry in cancer registration in Switzerland

Under the Cancer Registration Act, three entities have clearly defined roles (**Fig. 1**): the Childhood Cancer Registry (ChCR), Cantonal Cancer Registries (CCRs), and the National Agency for Cancer Registration (NACR). Healthcare providers are legally obliged to report cancer diagnoses, treatments, and follow-up information to the appropriate registry.

**Figure 1:**
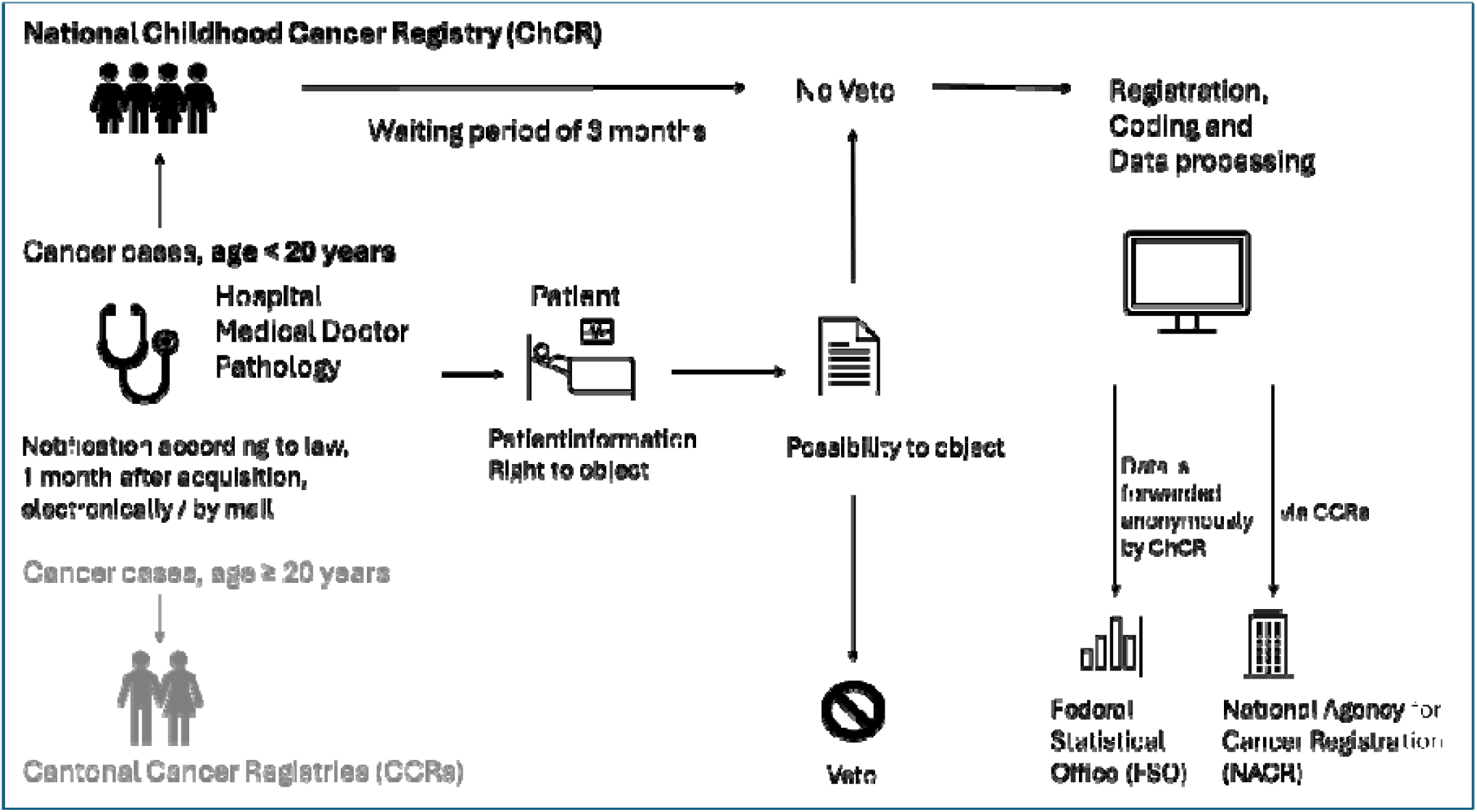
Process for recording cancer cases in children and adolescents in the Swiss Childhood Cancer Registry since the Cancer Registration Act (2020). Abbreviations: CCR, Cantonal Cancer Registry; ChCR, Childhood Cancer Registry; FSO, Federals Statistical Office; NACR, National Agency for Cancer Registration.

The ChCR (www.childhoodcancerregistry.ch) centrally records all cancer cases diagnosed in individuals under 20 years of age across Switzerland. Since 2020, it receives data from a broad range of reporting sources—not only from specialized paediatric oncology clinics, though they account for the majority of cases. This has improved completeness, particularly among adolescents aged 16 and older, who are sometimes treated outside pediatric settings. To enable coverage of all age groups in the cantonal registries, the ChCR provides each responsible cantonal registry with a copy of its data. The role and tasks of the ChCR are described in detail on the national information platform Cancer Registration in Switzerland [10].

*Cantonal Cancer Registries (CCRs)* are responsible for the initial registration of cancers in adults aged 20 and older within their regions, ensuring accurate and timely reporting by healthcare providers. The *National Agency for Cancer Registration (NACR)* coordinates and monitors data quality across the national cancer registration system, ensures standardized data collection, is responsible for data requests and statistical reporting for all ages. For childhood and adolescent cancers, the ChCR collaborates with the NACR on data quality, routine cancer reporting and represents Switzerland in international collaborations.

### Inclusion criteria

The ChCR records all incident cancers diagnosed in children, adolescents, and young adults up to their 20th birthday who belong to the permanent resident population of Switzerland at the time of diagnosis. Eligible diagnoses include all malignant tumors (ICD-10 codes C00–C97); benign tumors of the central nervous system (CNS; D32, D33, D35); histiocytosis (D76); selected in situ neoplasms (D00–D03, D05–D09); neoplasms of uncertain or unknown behavior of all sites (D37–D48); aplastic anemia (D61); and certain rare non-malignant hematological diseases. Annex 1 of the Cancer Registration Ordinance defines reportable diagnoses [11][12].

Diagnostic coding follows the same international standards used for adult cancers, including the International Classification of Diseases (ICD-10), the International Classification of Diseases for Oncology (ICD-O-3) and the TNM classification. In addition, pediatric systems are applied, namely the International Classification of Childhood Cancer (ICCC-3) and the Toronto Childhood Cancer Staging Guidelines, which enable age-appropriate grouping and staging of childhood cancers.

### Database, registration, and quality control

In 2020, the ChCR migrated its database from the ISPM to a central system developed and hosted by the Federal Office of Information Technology, Systems and Telecommunication. CCRs did not adopt this system, which created challenges for data harmonization. Therefore, the ChCR is currently transitioning to NICERStat **–** the registration software used by the CCRs.

Following the initial reporting of a new case, the law mandates a three-month waiting period before registration is finalized, allowing time for patients or legal guardians to exercise their right to object (opt out). If no objection is received within this period, the case is officially registered. Quality control is carried out rigorously by comparing the data with other sources (annual hospital statistics, causes of death data from the FSO, municipality data via CCRs), manually checking all cases using the dual control principle, and automated plausibility checks, including the Quality Check Software developed by the European Network of Cancer Registries (ENCR). These procedures ensure consistency, accuracy, and completeness of the data.

### Collected information (national data dictionary of variables)

The ChCR collects **a core set of variables** identical to those collected by the CCRs for adults (basic variables) [13]. These variables include personal and demographic information such as sex, date of birth, nationality, and place of residence at diagnosis, as well as detailed diagnostic data, such as date of diagnosis, tumor site, morphology, and stage. Additionally, the variables include the first course of treatment. Treatment details cover surgery, chemotherapy, radiotherapy, hematopoietic stem cell transplantation, and other modalities. The registry also collects longitudinal data on disease progression, relapses, second malignancies, and survival status.

In addition to the standard dataset, the ChCR records **supplementary variables** not routinely collected by most adult tumors [14]. These include information on additional treatment courses, cancer predisposition syndromes, and extended follow-up.

The ChCR annually obtains detailed causes of death data on the Swiss resident population aged 0-19 years from FSO. These data form the basis for calculating cancer specific mortality and allow the ChCR to identify potentially missed cancer cases (death certificate notifications). Causes of death are coded according to International Statistical Classification of Diseases and Related Health Problems (ICD version 10; version 9 for deaths before 1995). Overall, the ChCR dataset mirrors that of the former Swiss Childhood Cancer Registry (SCCR) before the implementation of the Cancer Registration Act, maintaining continuity while adapting to new legal and technical frameworks.

### Confidentiality and data protection

Mandated by the Federal Office of Public Health, the ChCR is the only national cancer registry in Switzerland. Research using data from the ChCR falls under the Human Research Act and the Swiss Data Protection Law, and is aligned with the European General Data Protection Regulation (GDPR). The handling of these sensitive data falls under the authority of the Federal Data Protection and Information Commissioner (EDÖB, Eidgenössischer Datenschutz- und Öffentlichkeitsbeauftragter).

### Ethics

In 2004, the former SCCR received a special registry authorization (Sonderbewilligung) from the Swiss Federal Commission of Experts for Professional Secrecy in Medical Research. This was replaced in June 2007 by a general authorization (Generelle Bewilligung), which permitted nationwide collection of childhood and adolescent cancer data with written, oral or silent consent. After the Human Research Act came into force in 2014, the registry obtained ethical approval from the ethics committee of the Canton of Bern (166/2014). Since 2020, ethical approval is no longer required, as patients’ rights are explicitly defined by law as; 1) the right to be informed about the registration; 2) the right to access their data; and 3) the right to object to registration (opt out). However, research projects that want to use registry data require ethics approval.

### Data availability

Individual patient data from the ChCR are available for research under strict conditions. Researchers must submit a detailed project proposal and obtain approval from a cantonal ethics committee [12]. All data sharing must comply with Swiss data protection laws, and data are pseudonymized or anonymized. A data transfer and usage agreement is required to ensure confidentiality and proper handling. Access to data is granted only to scientifically sound projects that align with the goals of the Cancer Registration Act, upon approval of the ChCR management board. Arrangements are currently underway to provide data through a secure platform, where researchers will be able to access but not export the data (remote analysis environment).

### Statistical analysis

The ChCR follows the common statistical procedures for routine reporting on cancer incidence, mortality, survival, and prevalence. These have been nationally harmonized between the ChCR, NKRS, and the BfS [15]. In this paper, we present data on incidence, mortality, and survival based on the ChCR data from 2014-2023, the most recent 10-year period with completed registration.

We calculated incidence and mortality rates as the number of new primary tumors or cancer deaths divided by person-years at risk, based on population data from the Swiss FSO [16]. For children (0-15), we included all cases recorded in the registry. For adolescents (16–19 years), nationwide coverage has only been complete since 2020, while before that, registration of cases not treated in SPOG centers relied mainly on linkage with existing cantonal registries; therefore, we extrapolated incidence counts for 2014–2019 to the national level based on cases from cantons with existing registries, applying population-based weights according to standard procedures [15]. Mortality data require no extrapolation as they are based on mandatory reporting and central recording of deaths by the FSO and can be considered complete for the entire country. Rates are reported per 1 million person-years and age-standardized to the 2013 European Standard Population [17], with 95% confidence intervals derived using the method of Fay and Feuer, based on the gamma distribution and accounting for the upweighting of incident adolescent cases [15]. We calculated observed survival for 1–10 years after diagnosis using Kaplan–Meier estimation and the period approach [15]. This approach considers only person years at risk that fall into a recent observation window (here 2014-2023) allowing delayed entry and right censoring and thus ensuring that estimates reflect the contemporary follow-up experience. We also report proportions of ICCC-3 main diagnostic groups (plus Langerhans cell histiocytosis) relative to all cases diagnosed between 2014 and 2023. We used R version 4.5.0 for all analyses. Full methodological details are described in *Statistical Methods for Cancer Reporting in Switzerland* [15].

## Results

### Overall dataset included in the registry

Since the beginning of childhood cancer registration in Switzerland in 1976 until December 31, 2023, the registry recorded 12,665 cancer cases that fulfilled the revised inclusion criteria, diagnosed among 12,479 patients. Absolute numbers of incident cases increased over time, mainly due to population growth in Switzerland and improvements in cancer registration (**Fig.2**). Given the relatively low case counts, year-to-year fluctuations are considerable.

**Figure 2:**
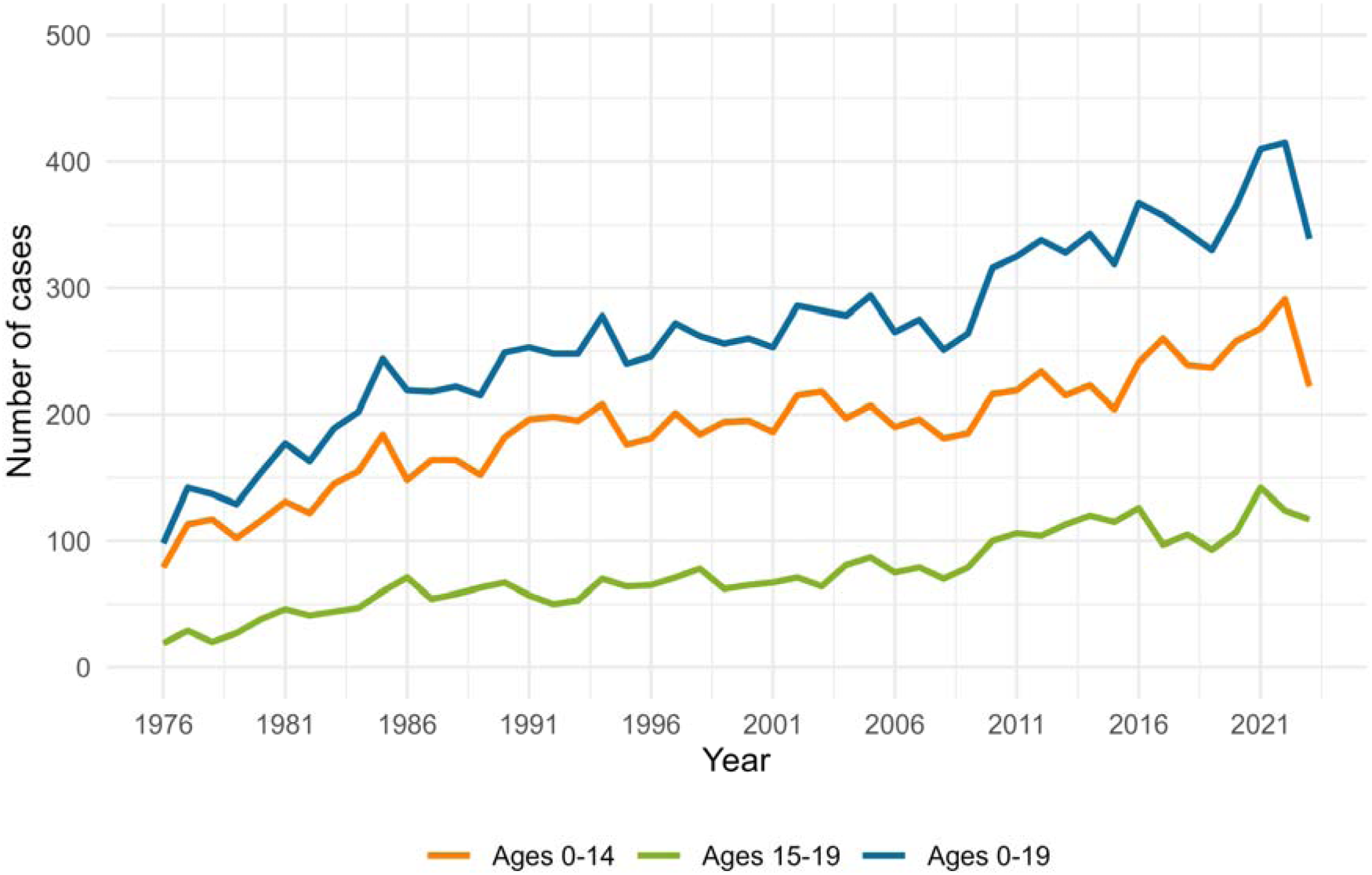
Annual number of cancer cases registered by the ChCR among children (0-14 years) and adolescents (15-19 years) living in Switzerland at time of diagnosis (1976-2023, ICCC-3 main groups and Langerhans cell histiocytosis). Abbreviations: ChCR, Childhood Cancer Registry; ICCC-3, International Classification of Childhood Cancer, third edition.

Between 2014 and 2023, 2,441 new cases were diagnosed among children (0–14 years) and an estimated 1,214 cases (1,146 registered cases) among adolescents (15–19 years). The distribution of cancer types varied substantially by age (**Fig.3**). In infants, embryonal tumors were most frequent, including neuroblastoma (22%), retinoblastoma (11%), kidney tumors (7%) and liver tumors (4%). Brain and other CNS tumors accounted for 17% of cases in infants and increased to 27% at ages 1–4 and 31% at 5–9, before declining to 26% at 10–14 and 18% at 15–19 years.

**Figure 3:**
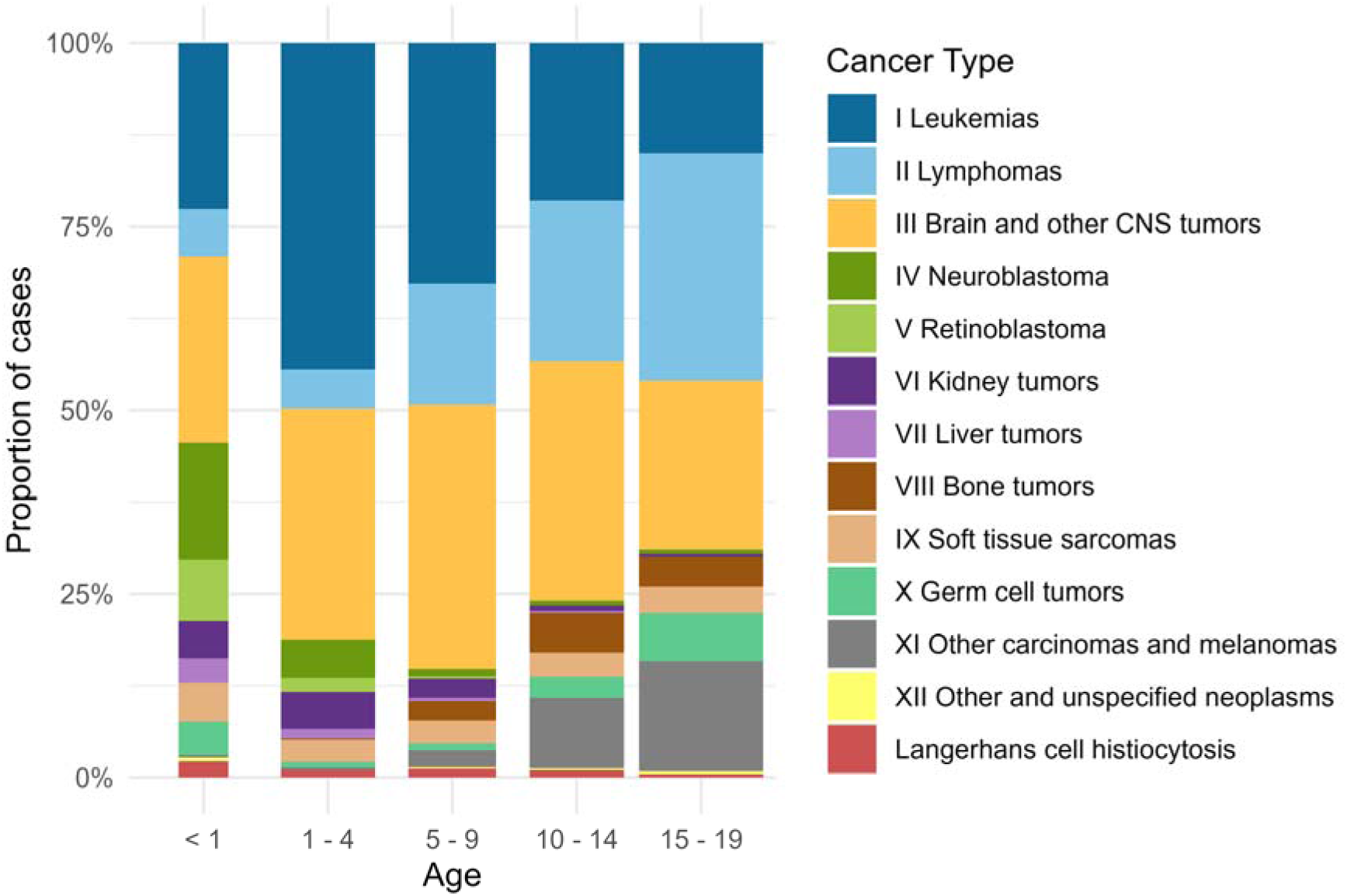
Proportions of different cancer types from infancy to age 19 years, by age at diagnosis (2014-2023, ICCC-3 main groups and Langerhans cell histiocytosis). The width of each bar is proportional to the number of cases in the age group, so wider bars indicate more diagnosed cases. Abbreviations: CNS, central nervous system; ICCC-3, International Classification of Childhood Cancer, third edition.

Leukemia, mainly acute lymphoblastic leukemia (ALL), was the leading cancer type in early childhood, peaking at 37% in children aged 1–4. Their share declined to 29% at 5–9 years, 17% at 10–14 years and 11% in adolescents. By contrast, tumor types that were rare in early childhood became more common at older ages: lymphomas accounted for 18% of cases at 10– 14 years and 24% at 15–19 years; carcinomas and melanomas for 16% and 23%, respectively; bone tumors for 9% and 6%. Germ cell tumors were relatively common in infants (6%) and again at ages 15–19 years (10%). Soft tissue sarcomas were less frequent overall but stable across ages, representing 7% in the first year of life and 5% in the other age groups. Overall, in children the most frequent diagnostic groups were CNS tumors (27%), leukemias (26%) and lymphomas (11%), while in adolescents lymphomas (24%), carcinomas and melanomas (23%) and CNS tumors (18%) predominated (**Tables 1 and 2**).

**Table 1:**
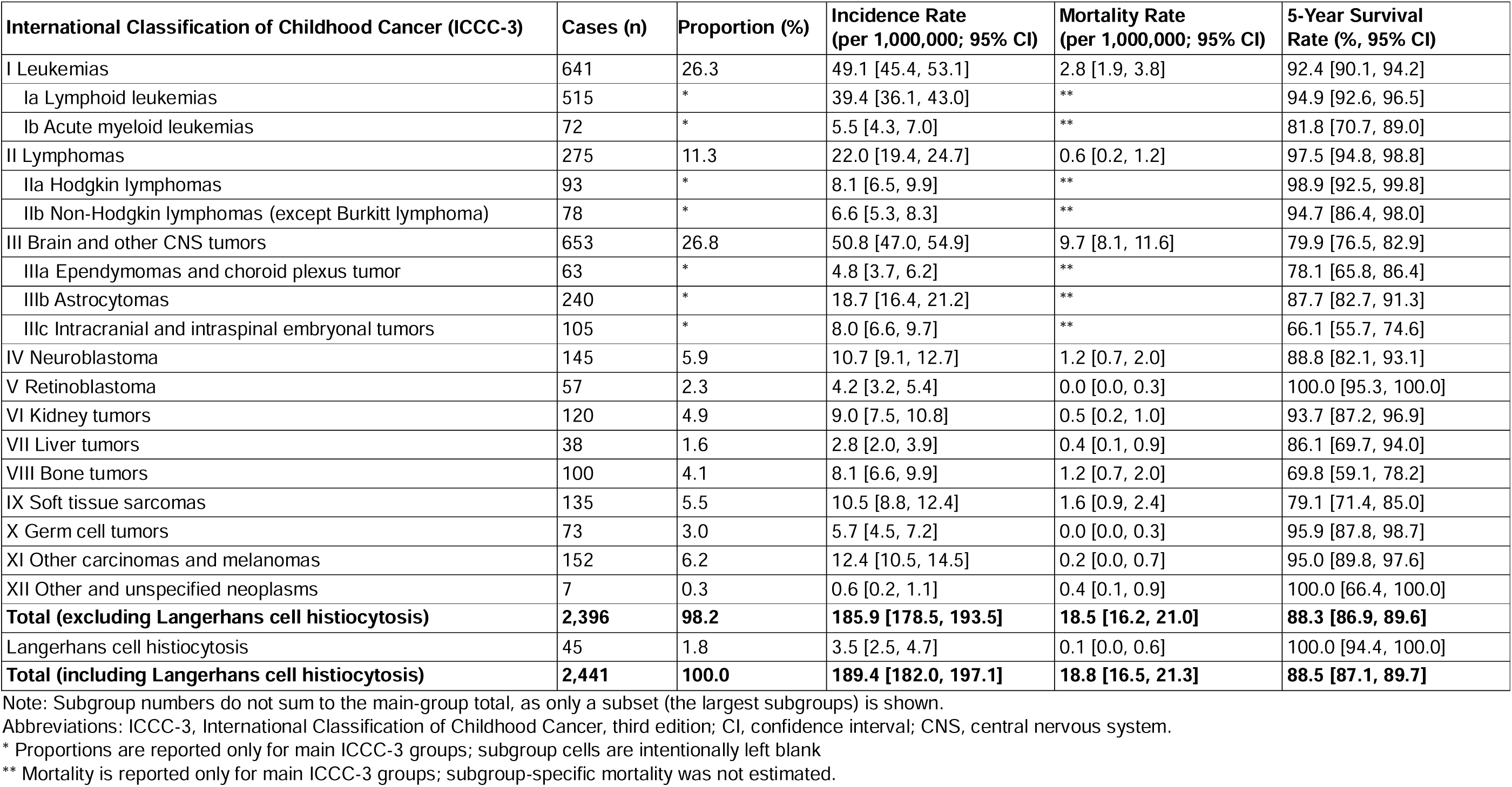
Ages 0–14, 2014–2023: ICCC-3 diagnostic groups and subgroups with estimated case numbers, proportions, average annual standardized incidence and mortality rates per 1,000,000 person-years (95% CI), and 5-year survival (95% CI); both sexes combined.

| International Classification of Childhood Cancer (ICCC-3) | Cases (n) | Proportion (%) | Incidence Rate<br>(per 1,000,000; 95% CI) | Mortality Rate<br>(per 1,000,000; 95% CI) | 5-Year Survival<br>Rate (%; 95% CI) |
| --- | --- | --- | --- | --- | --- |
| I Leukemias | 641 | 26.3 | 49.1 [45.4, 53.1] | 2.8 [1.9, 3.8] | 92.4 [90.1, 94.2] |
| Ia Lymphoid leukemias | 515 | * | 39.4 [36.1, 43.0] | ** | 94.9 [92.6, 96.5] |
| Ib Acute myeloid leukemias | 72 | * | 5.5 [4.3, 7.0] | ** | 81.8 [70.7, 89.0] |
| II Lymphomas | 275 | 11.3 | 22.0 [19.4, 24.7] | 0.6 [0.2, 1.2] | 97.5 [94.8, 98.8] |
| IIa Hodgkin lymphomas | 93 | * | 8.1 [6.5, 9.9] | ** | 98.9 [92.5, 99.8] |
| IIb Non-Hodgkin lymphomas (except Burkitt lymphoma) | 78 | * | 6.6 [5.3, 8.3] | ** | 94.7 [86.4, 98.0] |
| III Brain and other CNS tumors | 653 | 26.8 | 50.8 [47.0, 54.9] | 9.7 [8.1, 11.6] | 79.9 [76.5, 82.9] |
| IIIa Ependymomas and choroid plexus tumor | 63 | * | 4.8 [3.7, 6.2] | ** | 78.1 [65.8, 86.4] |
| IIIb Astrocytomas | 240 | * | 18.7 [16.4, 21.2] | ** | 87.7 [82.7, 91.3] |
| IIIc Intracranial and intraspinal embryonal tumors | 105 | * | 8.0 [6.6, 9.7] | ** | 66.1 [55.7, 74.6] |
| IV Neuroblastoma | 145 | 5.9 | 10.7 [9.1, 12.7] | 1.2 [0.7, 2.0] | 88.8 [82.1, 93.1] |
| V Retinoblastoma | 57 | 2.3 | 4.2 [3.2, 5.4] | 0.0 [0.0, 0.3] | 100.0 [95.3, 100.0] |
| VI Kidney tumors | 120 | 4.9 | 9.0 [7.5, 10.8] | 0.5 [0.2, 1.0] | 93.7 [87.2, 96.9] |
| VII Liver tumors | 38 | 1.6 | 2.8 [2.0, 3.9] | 0.4 [0.1, 0.9] | 86.1 [69.7, 94.0] |
| VIII Bone tumors | 100 | 4.1 | 8.1 [6.6, 9.9] | 1.2 [0.7, 2.0] | 69.8 [59.1, 78.2] |
| IX Soft tissue sarcomas | 135 | 5.5 | 10.5 [8.8, 12.4] | 1.6 [0.9, 2.4] | 79.1 [71.4, 85.0] |
| X Germ cell tumors | 73 | 3.0 | 5.7 [4.5, 7.2] | 0.0 [0.0, 0.3] | 95.9 [87.8, 98.7] |
| XI Other carcinomas and melanomas | 152 | 6.2 | 12.4 [10.5, 14.5] | 0.2 [0.0, 0.7] | 95.0 [89.8, 97.6] |
| XII Other and unspecified neoplasms | 7 | 0.3 | 0.6 [0.2, 1.1] | 0.4 [0.1, 0.9] | 100.0 [66.4, 100.0] |
| **Total (excluding Langerhans cell histiocytosis)** | **2,396** | **98.2** | **185.9 [178.5, 193.5]** | **18.5 [16.2, 21.0]** | **88.3 [86.9, 89.6]** |
| Langerhans cell histiocytosis | 45 | 1.8 | 3.5 [2.5, 4.7] | 0.1 [0.0, 0.6] | 100.0 [94.4, 100.0] |
| **Total (including Langerhans cell histiocytosis)** | **2,441** | **100.0** | **189.4 [182.0, 197.1]** | **18.8 [16.5, 21.3]** | **88.5 [87.1, 89.7]** |

Note: Subgroup numbers do not sum to the main-group total, as only a subset (the largest subgroups) is shown.
Abbreviations: ICCC-3, International Classification of Childhood Cancer, third edition; CI, confidence interval; CNS, central nervous system.
\* Proportions are reported only for main ICCC-3 groups; subgroup cells are intentionally left blank
\*\* Mortality is reported only for main ICCC-3 groups; subgroup-specific mortality was not estimated.

**Table 2:** Ages 15–19, 2014–2023: ICCC-3 diagnostic groups and subgroups with estimated case numbers, proportions, average annual standardized incidence and mortality rates per 1,000,000 person-years (95% CI), and 5-year survival (95% CI); both sexes combined.

| International Classification of Childhood Cancer (ICCC-3) | Cases (n) | Proportion (%) | Incidence Rate<br>(per 1,000,000; 95% CI) | Mortality Rate<br>(per 1,000,000; 95% CI) | 5-Year Survival<br>Rate (%; 95% CI) |
| --- | --- | --- | --- | --- | --- |
| I Leukemias | 140 | 11.5 | 32.4 [26.9, 38.6] | 4.9 [3.0, 7.5] | 89.0 [82.2, 93.4] |
| Ia Lymphoid leukemias | 73 | * | 16.9 [13.0, 21.5] | ** | 90.5 [80.0, 95.6] |
| Ib Acute myeloid leukemias | 28 | * | 6.6 [4.3, 9.7] | ** | 77.2 [56.1, 89.0] |
| II Lymphomas | 287 | 23.6 | 66.6 [58.6, 75.4] | 2.3 [1.1, 4.3] | 95.3 [91.9, 97.3] |
| IIa Hodgkin lymphomas | 217 | * | 50.4 [43.5, 58.1] | ** | 97.4 [93.9, 98.9] |
| IIb Non-Hodgkin lymphomas (except Burkitt lymphoma) | 56 | * | 13.1 [9.7, 17.3] | ** | 85.0 [71.1, 92.6] |
| III Brain and other CNS tumors | 213 | 17.5 | 49.5 [42.6, 57.1] | 7.9 [5.5, 11.0] | 89.4 [83.8, 93.1] |
| IIIa Ependymomas and choroid plexus tumor | 18 | * | 4.2 [2.4, 6.8] | ** | 100.0 [84.6, 100.0] |
| IIIb Astrocytomas | 62 | * | 14.3 [10.8, 18.7] | ** | 84.4 [71.1, 92.0] |
| IIIc Intracranial and intraspinal embryonal tumors | 15 | * | 3.4 [1.8, 5.8] | ** | 83.3 [48.2, 95.6] |
| IV Neuroblastoma | 10 | 0.8 | 2.4 [1.1, 4.5] | 0.9 [0.3, 2.4] | 85.7 [33.4, 97.9] |
| V Retinoblastoma | 0 | 0.0 | *** | *** | *** |
| VI Kidney tumors | 8 | 0.6 | 1.8 [0.7, 3.8] | 0.5 [0.1, 1.7] | 87.5 [38.7, 98.1] |
| VII Liver tumors | 0 | 0.0 | *** | *** | *** |
| VIII Bone tumors | 77 | 6.3 | 17.8 [13.8, 22.6] | 6.3 [4.1, 9.1] | 65.1 [52.9, 74.9] |
| IX Soft tissue sarcomas | 65 | 5.3 | 15.1 [11.4, 19.6] | 5.1 [3.2, 7.7] | 63.4 [50.9, 73.5] |
| X Germ cell tumors | 123 | 10.1 | 28.5 [23.3, 34.4] | 0.7 [0.1, 2.0] | 95.1 [89.5, 97.8] |
| XI Other carcinomas and melanomas | 277 | 22.8 | 64.4 [56.5, 73.1] | 1.4 [0.5, 3.0] | 95.6 [92.2, 97.5] |
| XII Other and unspecified neoplasms | 7 | 0.6 | 1.6 [0.6, 3.5] | 1.2 [0.4, 2.7] | 46.9 [9.0, 78.8] |
| **Total (excluding Langerhans cell histiocytosis)** | **1,206** | **99.3** | **280.0 [263.4, 297.4]** | **31.1 [26.1, 36.8]** | **89.7 [87.7, 91.4]** |
| Langerhans cell histiocytosis | 8 | 0.7 | 1.8 [0.7, 3.8] | 0.0 [0.0, 0.9] | 100.0 [63.1, 100.0] |
| **Total (including Langerhans cell histiocytosis)** | **1,214** | **100.0** | **281.8 [265.1, 299.3]** | **31.3 [26.3, 37.1]** | **89.7 [87.8, 91.4]** |

Note: Subgroup numbers do not sum to the main-group total, as only a subset (the largest subgroups) is shown.
Abbreviations: ICCC-3, International Classification of Childhood Cancer, third edition; CI, confidence interval; CNS, central nervous system.
\* Proportions are reported only for main ICCC-3 groups; subgroup cells are intentionally left blank
\*\* Mortality is reported only for main ICCC-3 groups; subgroup-specific mortality was not estimated.
\*\*\* No cases were observed in this subgroup during 2014–2023.

### Incidence and mortality

Between 2014 and 2023, the overall incidence per million person-years was 189 (95% CI 182-197) among children (184 for girls and for 194 for boys) and 282 (95% CI 265-299) for adolescents (279 for females and 284 for males). For both sexes, incidence rates were high in infancy, decreased during mid-childhood, and rose again during adolescence. Males tended to have higher incidence rates than females (**Fig. 4**).

**Figure 4:**
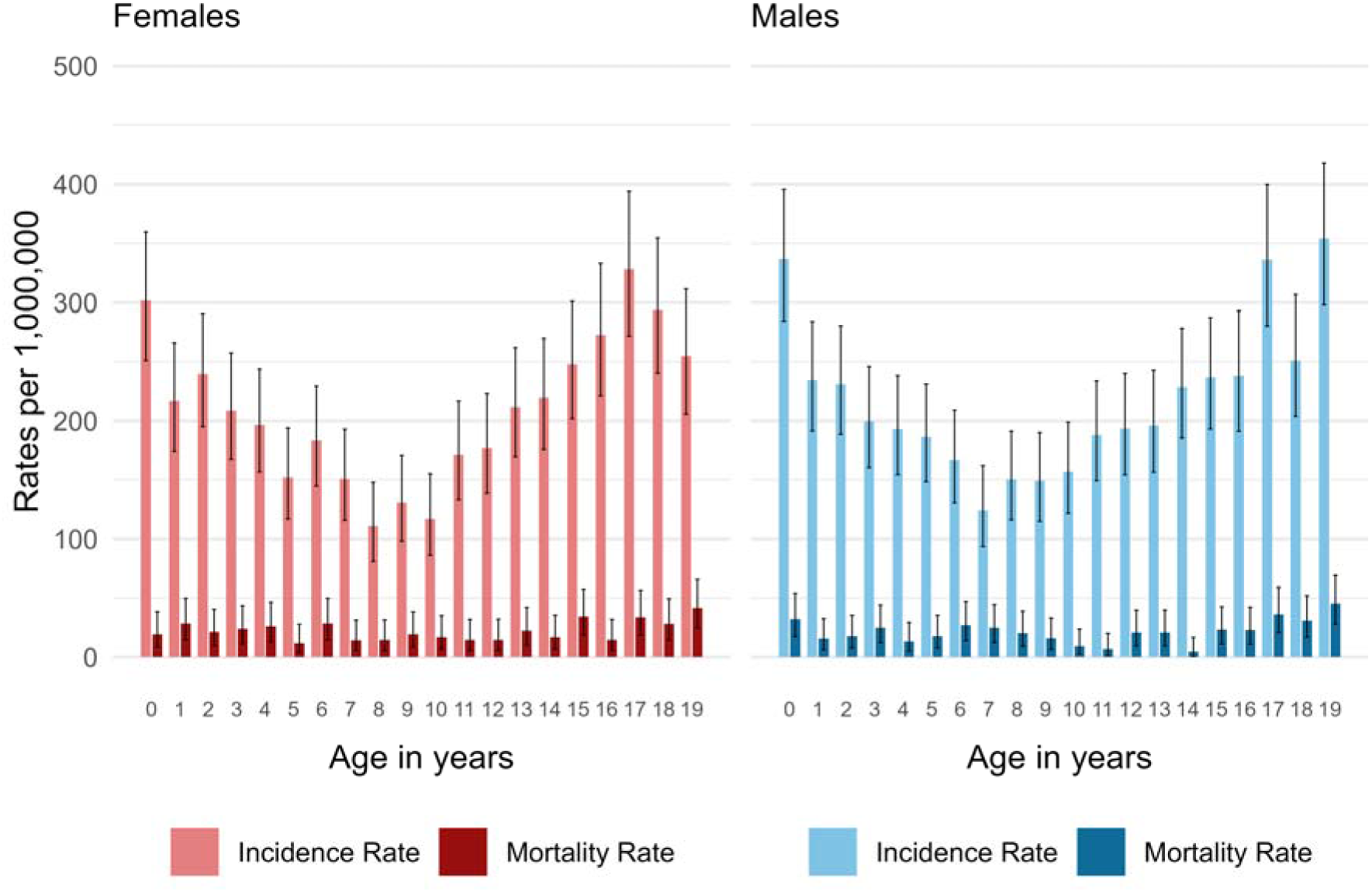
Age- and sex-specific average annual incidence and mortality rates (per 1 million person-years, 95% confidence intervals) in children and adolescents, 2014–2023, ICCC-3 main diagnostic groups and Langerhans cell histiocytosis. Abbreviations: ICCC-3, International Classification of Childhood Cancer, third edition.

Overall cancer mortality was low, with rates of 19 per million (95% CI 17–21) in children and 31 per million (95% CI 26–37) in adolescents (Fig. 4, Tables 1 and 2). In both age groups, mortality was highest for CNS tumors, with rates of 10 per million (95% CI 8–12) for children and 8 per million (95% CI 6–11) for adolescents. We could not estimate mortality rates for subgroups because re-classification from ICD to ICCC-3 of FSO causes of death data is only feasible at main group level.

### Survival

**Fig. 5** presents observed survival for cancers between 2014 and 2023. The overall 5-year survival was 89% (95% CI 87–90) in children and 90% (95% CI 88–91) in adolescents. In children, survival exceeded 90% for leukemia and lymphomas but was lower for CNS tumors (80%, 95% CI 77-83), bone tumors (70%, 95% CI 59-78), and soft tissue sarcomas (79%, 95% CI 71-85) (**Table 1**). Adolescents had slightly higher survival than children for CNS tumors (89%, 95% CI 84-93), while survival for other diagnostic groups was broadly similar (Table 2). Across all age groups, the highest survival rates occurred for lymphomas, germ cell tumors, carcinomas and melanomas (**Table 1 and 2**).

**Figure 5:**
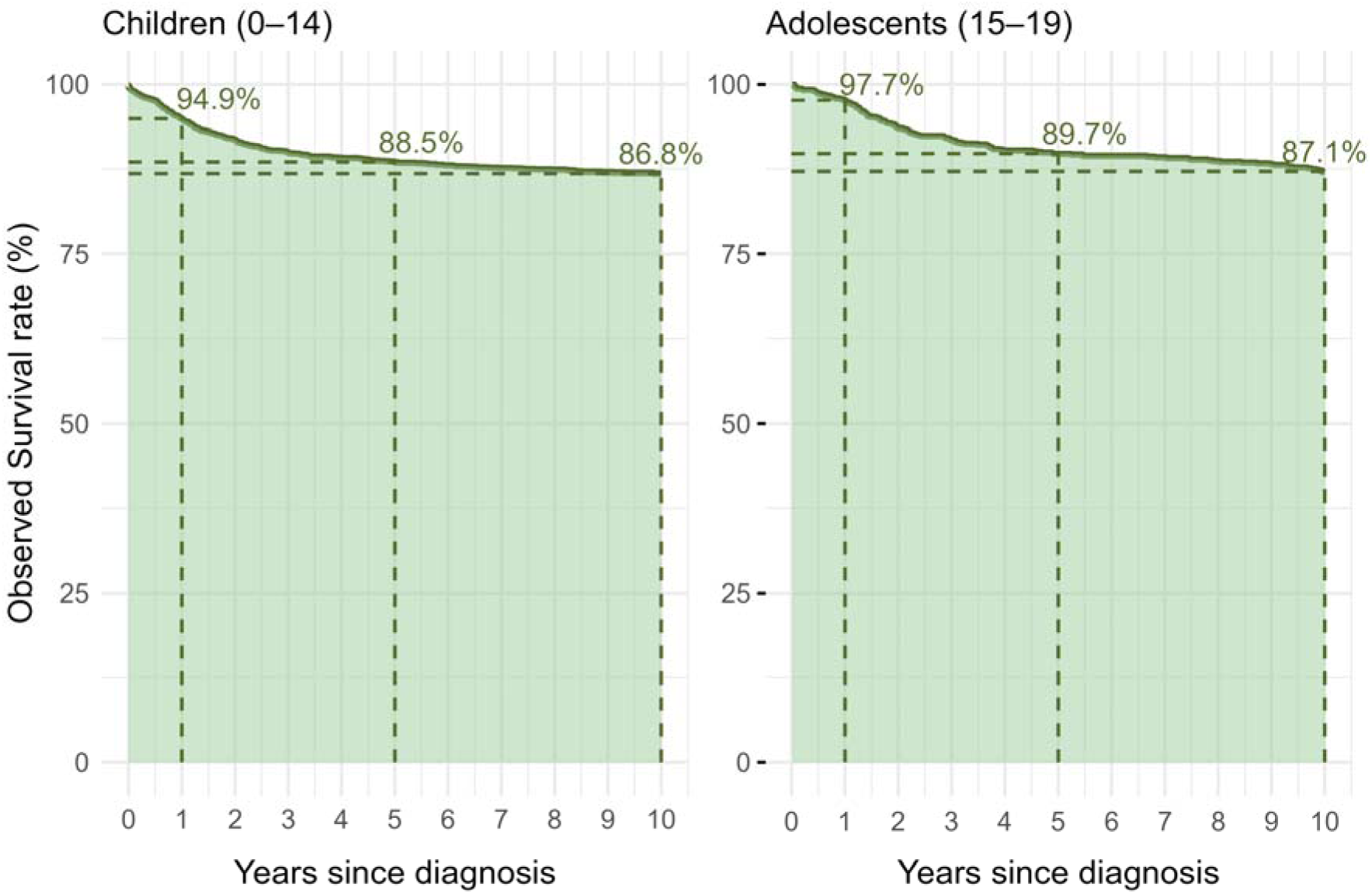
Observed Survival of children (0-14 years) and adolescents (15-19 years): rate over time after cancer diagnosis based on data from 2014–2023 (ICCC-3 main groups and Langerhans cell histiocytosis). Abbreviations: ICCC-3, International Classification of Childhood Cancer, third edition.

## Use of Childhood Cancer Registry Data for Research

Data from the ChCR have been widely used for research across multiple domains:

- **Cancer epidemiology**: We have described incidence and survival trends [18, 19], second primary neoplasms [20, 21], examined late mortality [22], and investigated associations between socioeconomic status and cancer incidence and outcomes [23, 24]. Current projects focus on the epidemiology of cancers among adolescents and young adults (AYA) [25].
- **Cancer registry methodology**: Studies have assessed data completeness and analyzed death certificate notifications [26, 27]. We are currently updating our methodological approaches to assess registration completeness and timeliness [28].
- **International collaboration**: ChCR data contribute regularly to international benchmarking studies, including CONCORD (cancer survival trends) [29], ENCR-ECIS (European cancer burden comparisons) [30], IARC projects such as CRICCS (secondary cancer risk) [31], BENCHISTA (stage-specific survival benchmarking) [32], and EUROCARE (European cancer outcomes) [33].
- **Clinical research**: ChCR data support clinical studies by identifying eligible cases or providing supplementary information, particularly in collaboration with pediatric oncology centers [20, 34–39].
- **Biobank collaboration**: ChCR data are used in joint studies with biobanks. This includes germline DNA biobanking in the national germline DNA Biobank for childhood cancer BISKIDS, which is part of the Geneva biobank BaHOP and hosted at the University of Geneva. Further data is used in conjunction with the tumor sample biobank SPHO, which is hosted at the University Children’s hospital Zurich. These datasets led to the study GECCOS which examines the influence of germline DNA variants on complications after childhood cancer [20].
- **Etiological research**: Studies have examined spatial and temporal cancer clustering and examined associations with environmental exposures such as ionizing/non-ionizing radiation, air pollution, and population mixing [40–45].
- **Survivorship research**: The Swiss Childhood Cancer Survivor Study (SCCSS, www.swiss-ccss.ch) follows all five-year survivors through repeated questionnaires. Research has addressed late effects (pulmonary, cardiac, auditory, mental health), health behaviors, social outcomes (education, partnerships, income), and parental impact and quality of follow-up care [46–66]. Nested studies conduct clinical assessments like lung function tests, echocardiography, and hearing exams [38, 39, 67].

A comprehensive list of publications using data from the ChCR is available on the registry’s website.

## Discussion

This study describes the development of childhood cancer registration in Switzerland and presents recent data on incidence, mortality, and survival. What began in 1976 as a voluntary, registry run by pediatric oncologists evolved into the Swiss Childhood Cancer Registry (SCCR) and is now, under the Cancer Registration Act, the federally mandated Childhood Cancer Registry (ChCR). The ChCR has become a comprehensive and complete national database and a critical resource for research. More than 200 scientific publications have used data from the ChCR.

### Comparison to other childhood cancer registries

Our results align with data from other high-income countries. Incidence rates, cancer type distribution, and survival are broadly comparable with childhood cancer registry statistics in neighboring countries, although direct comparisons are often limited by differences in time periods covered, inclusion criteria, age-ranges, and reference populations used for age-standardization. In Switzerland, we observed a cancer incidence of 186 per million for children aged ≤14 years (2014–2023, excluding Langerhans cell histiocytosis). By comparison, the French national registry of childhood cancers (RNCE) reported an incidence of 161 per million for children ≤15-years (2014–2020, 1976 European standard population used [68]), and the German Childhood Cancer Registry (GCCR) reported 175 per million for children and adolescents <18 years (Segi’s world standard population used [69]). As for other registries, the annual number of incident cases in ChCR per year has increased, reflecting both population growth and improvements in cancer reporting. Our previous work including data up to 2014 suggested a possible real increase in the incidence of leukemia and brain tumors [18]. Given the high rates reported for CNS tumors in this study, an in-depth investigation of trends and potential drivers, using the most recent data, is planned.

Childhood cancer survival rates in Switzerland are among the highest worldwide [29]. Based on follow-up data during 2014–2023, ten-year survival for both children and adolescents reached 87% (**Fig. 5**). We found socioeconomic disparities for children diagnosed with cancer between 1991-2006 in survival, particularly for brain tumors [23]. We plan to reexamine these using the current dataset, to understand measures needed to ensure optimal survival for all patients.

### Strenghts and limitations

The transition from the voluntarily maintained Swiss Childhood Cancer Registry (SCCR) to the current federally mandated ChCR led to substantial improvements. Securing the continuation of decades of childhood cancer registration with a high completeness enables seamless monitoring and investigation of trends. Adolescents aged 16–19, previously underrepresented when treated outside pediatric centers, are now systematically captured. Cases reported only by pathology laboratories, which may have been missed under the previous clinician-led model, are now included, further improving completeness. The Cancer Registration Act created a transparent legal framework, mandating standardized and complete reporting and giving patients the right to opt out. Standardized comparison procedures using hospital discharge data and mortality statistics support systematic validation of completeness and survival estimates, thereby providing stronger quality control. Stable federal funding secures long-term sustainability, independent of research grants or donations. These changes strengthen the ChCR’s reliability and public trust.

The new framework also introduced limitations. Data availability is delayed by a three-month waiting period and complex data exchanges between ChCR, CCRs and the NACR. Coordination gaps and database incompatibilities between these registries hinder linkage of second malignancies to the same patient, delay timely monitoring of incidence trends and make monitoring and research on late effects more difficult. Additional data protection requirements and new access protocols, while essential for confidentiality, complicate data use. Together, these limitations reduce timeliness and clinical utility.

### Outlook

Several key developments could further strengthen childhood cancer registration in Switzerland. Completeness among adolescents is a priority, including dedicated analyses to identify and address gaps or inconsistencies. A unified national cancer database integrating data from all cancer registries, including the ChCR, would facilitate case linkage to identify second primary cancers and reduce delays. Direct data transfers from clinical data warehouses and electronic health records could further improve data quality and completeness, enhance timeliness, and reduce administrative workload. Eliminating the current three-month waiting period for registration would accelerate reporting without compromising the patient’s right to object, because patients can request anonymization of their records at any time.

### Conclusion

The ChCR has developed into a long-standing, high-quality, and robust population-based registry. Mandatory reporting, strong national collaboration, and a federal framework ensure high completeness and sustainable operation. The ChCR provides a critical resource for both epidemiological and clinical research on childhood and adolescent cancers and allows Switzerland to contribute to international research initiatives. Future efforts should focus on improving timeliness, enabling direct transfer of clinical data from hospital records and linkage across registries, as well as promoting the responsible use of registry data. These steps will further strengthen the ChCR as a foundation for cancer surveillance, research, and policy.

## Statements & Declarations

## Acknowledgements

We thank all patients for their trust in the Childhood Cancer Registry. We thank Nicolas Weid for his sustained scientific and clinical input to the Swiss Childhood Cancer Registry and Eleftheria Michalopoulou for her previous statistical contributions.

## Author contribution

GS: data management, writing — review and editing; MAH, BS: formal analysis, writing — visualization, review and editing; SdV, CCK and UK: describing and visualizing of the ChCR operative functioning, writing — review and editing; CS, FNB, NW: writing — review and editing; CEK: conceptualization, methodology, first draft, writing — review and editing, funding acquisition, supervision. All authors read and approved of the final manuscript.

## Data availability

Researchers interested in collaborative work can contact the data request team of the ChCR to discuss planned projects or analyses of existing data. Data request forms can be found on our website: (https://www.childhoodcancerregistry.ch/data/data-requests) [12].

## Ethics approval

A clear legal framework, defined in the Cancer Registration Act, mandates reporting and includes a transparent opt out process for patients, enhancing public trust and legal consistency, hence no ethical approval is needed for cancer registration in Switzerland.

## Consent to participate

Due to the patients’ right to object to the registration of cancer cases, no informed consent for registration is required since the Cancer Registration Act came into force.

## Consent to publish

Not applicable.

